# Assessing risk perceptions that contribute to tetanus toxoid maternal vaccine hesitancy in Kilifi County, Kenya

**DOI:** 10.1101/2021.04.11.21255279

**Authors:** Patience Kerubo Kiyuka, Rodgers Onsomu Moindi, Nickson Murunga, Naomi Muinga, Meshack Nzesei Mutua, Stanley Wanjala, Sheba Sandra, Noni Mumba, Evans Otieno Odhiambo, Lillian Mutengu, Halimu Suleiman Shauri

## Abstract

Vaccination is one of the most effective public health interventions today. However, there is a growing number of people who perceive vaccines as unsafe and unnecessary. Waning vaccine confidence threatens global immunization programmes, contributing to decreased immunization rates and outbreaks of vaccine-preventable diseases. We used a mixed-method approach surveying men and women of reproductive age and Focus Group Discussions with expectant mothers to understand maternal vaccine hesitancy within a rural setting of Kilifi County, Kenya. Of the 104 people surveyed, 70% of the participants were aware of the vaccine that expectant women receive and 26% stating that they know people in their community who have refused or were hesitant to take maternal vaccination. Reasons given for refusals include religion and rumors that have spread in the community that the tetanus toxoid vaccine was a family planning method. Stockout of the vaccine was identified as one of the healthcare factors affecting vaccine uptake. The majority of the respondents (84%) reported that they mainly trust a healthcare worker for vaccination information. Approximately 53% and 23% reported that they strongly agree or agreed, respectively, that vaccines for expectant women should be made compulsory. The availability of safe and effective maternal vaccines will only be beneficial if mothers choose to use them. Dissemination of accurate information and continuous engagement with the community members can build trust and confidence in vaccines.

## Introduction

Globally, the World Health Organization (WHO) estimates that in 2019 alone, 2.4 million children died in their first month of life (WHO 2020). The majority of these deaths were a result of infectious diseases. Although substantial progress has been made in child survival since 1990, neonatal deaths remain relatively high in sub-Saharan Africa compared to other regions. Children born in sub-Saharan Africa are ten times more likely to die in their first month of life than their counterparts in high-income countries (WHO 2020). Maternal immunization has the potential to contribute to a significant reduction in neonatal morbidity and mortality. For instance, it estimated that maternal tetanus vaccine reduces mortality from neonatal tetanus by 94% [95% confidence interval (CI) 80-98%] (Blencowe et al., 2010). Immunization of pregnant women offers a chance to transplacentally transfer protective antibodies to the fetus, conferring protection to the vulnerable infants in the first six months of life where they are exposed but have not developed adequate functional antibody response (Marshall et al., 2016).

Despite the compelling benefits achieved through vaccination, there is a growing concern about vaccine hesitancy. The WHO Strategic Advisory Group of Expert on Immunization defines vaccine hesitancy as ‘delay in acceptance or refusal of vaccination despite the availability of vaccination services’, which varies across time, place and the vaccine. Vaccine hesitant individuals may accept all vaccines but remain concerned about vaccines, some may refuse or delay some vaccines, but accept others; some individuals may refuse all vaccines.(Larson et al., 2014, 2015). In fact, WHO in 2019 listed vaccine hesitancy among the top ten public health threats that needs attention; a rallying call to governments, public health officials and advocacy groups to pay attention to this issue. Left unchecked, vaccine hesitancy and misinformation that can spread rapidly through the internet (Cornwall 2020; Enserink 2020), can significantly contribute to decrease in immunization rates and hamper the uptake of newly introduced vaccines (Schaetti et al., 2009; Larson et al., 2010; Dubé et al., 2015). For example in North East Nigeria, vaccine myths, hesitancy and misinformation that spread within the communities provided significant setbacks to the campaign to eradicate polio disease (Usman et al., 2019).

Currently recommended vaccines for pregnant women include: tetanus toxoid, influenza vaccine and Tdap vaccine (Larson Williams et al., 2019), although countries have different guidelines and can choose which vaccines to administer. The Kenya Expanded Programme on Immunization recommends that expectant women receive tetanus toxoid vaccine, although the Ministry of Health is exploring evidence based strategies ahead of the anticipation to roll out influenza vaccine (McMorrow et al., 2017; Dawa et al., 2019). The success of these vaccines depends on whether mothers decide to use them.

Several factors that contribute to vaccine hesitancy have been reported. These include: expectant women’s levels of knowledge and attitude towards the safety and efficacy of the vaccine (Eppes et al. 2013; Bushar et al. 2017), how the healthcare worker interacts with mothers (Bukenya and Freeman 1991) and health systems factors such as health care practices and vaccine logistical issues (Sridhar et al. 2014). Literature specifically looking at maternal vaccination in Kenya is sparse. Most studies have focused on coverage and implementation of Antenatal Care (ANC), and on whether ANC services contribute to better neonatal care and assessment of ANC services as a package (Arunda et al. 2017; Afulani et al. 2019). In this study, we aimed to assess factors contributing to maternal vaccine hesitancy withing a rural setting of Kilifi County in Kenya.

## Methods

### Study location

The study was conducted at Kilifi County, located along the Kenyan Coast. In Kenya, health delivery system is currently a devolved function of the 47 counties. It is organized into six levels but anticipated to transition to four levels: level four, National referral hospitals; level three, county hospitals; level two, primary care facilities (formerly the dispensaries and health centres); and level 1, community hospital (Muinga et al. 2020).

### Study population

Our study randomly sampled expectant women attending ANCs at Kilifi County Hospital (KCH), a level three hospital, and two dispensaries serving KCH, Mnarani and Ngerenya dispensaries, which are in level two. We also randomly selected men and women of reproductive age at the community level in four different locations/wards (Tezo, Chasimba, Kibarani and Sokoni) using Community Health Extension Officers. Selection of study location was based on immunization coverage from the Kilifi County’s Kenya Health Information System (KHIS). According to the 2019 KHIS data, the percentage of women who received first tetanus toxoid vaccine ranged from 6% to 61.2 % (lowest to highest) across the 35 administrative wards. We split the percentage in tetanus toxoid vaccine coverage into four quartiles (0-25, 25-50, 50-75 and 75-100) and then selected wards from each quartile based on ease of accessibility.

### Questionnaire and question guide design

Data collection question guides and questionnaire design was based on the 2012 guidelines provided by the WHO Strategic Advisory Group on Experts on immunization to assess vaccine hesitancy (Larson et al. 2015). The questionnaire had a mix of open-ended questions and closed questions on a five-point Likert scale. A technical team of senior scientists reviewed the questionnaire and question guides and modified appropriately to fit into the local context. The questions were translated into Kiswahili, a local language for ease of administration. Interview guides were used flexibly to support rich and broader discussion around maternal vaccine hesitancy issues.

### Data collection

Prior to data collection the study team underwent training on the overall aims and objectives of the study, the consenting process and the data quality and management processes. Following the training, we conducted four Focus Group Discussions (FGDs) with the expectant women and administered survey questionnaires to men and women of reproductive age (n=104). We also conducted interviews with six key informants. Data collection was done between September to October 2020.

### Data analysis

Quantitative survey data was analyzed using STATA version 16 and the discussion data analyzed using thematic analysis. Baseline characteristics of participants including gender, marital status, level of education, and religion are presented as frequencies and percentages. Maternal vaccine knowledge and reasons for refusal to take vaccine are presented as pie charts. Bar charts were used to present barriers to maternal tetanus toxoid vaccine uptake and Trust in healthcare workers and the government.

## Ethics statement

We obtained scientific and ethical approval from the Pwani University Ethics Review Committee; ERC/PU-STAFF/002/2020. The study got approval from the National Commission for Science Technology and Innovation (NACOSTI) under license number NACOSTI/P/20/6193. The county government department of health services authorized the study to be carried out in Kilifi County. Since study activities were conducted during the COVID-19 pandemic, the study team followed the infection control guidelines put in place by County Department of Health Services to reduce SARS-CoV-2 infections such as: social distancing; regular hand washing; use of personal protective equipment to protect oneself and participants; and measuring temperatures of study team before each field visits. Eligible participants gave written consent prior to participation in the study.

## Results

Survey respondents represented a random sample of 104 adults. Their characteristics and summary are shown in Table 1. Women were 63 (61%) of the study population and 80 (77%) of the participants were married. Most participants had primary level education. The median age was 33 years.

**Table 1.** Baseline characteristics for the study participants.

| Characteristics | n (%) |
| --- | --- |
| Gender |  |
| Male | 40 (38.46) |
| Female | 63 (60.58) |
| Missing | 1 (0.96) |
| Marital status |  |
| Single | 18 (17.31) |
| Married | 80 (76.92) |
| Divorced | 4 (3.85) |
| Missing | 2 (1.92) |
| Level of education |  |
| Never went to school | 7 (6.73) |
| Primary education | 46 (44.23) |
| Secondary education | 33 (31.73) |
| Diploma | 11 (10.58) |
| University education | 7 (6.73) |
| Religion |  |
| Christianity | 89 (85.58) |
| Islam | 12 (11.54) |
| Other | 1 (0.96) |
| Missing | 2 (1.92) |
| Age (in years) |  |
| Median Interquartile range (IQR) | 33 (27-43) |

### General vaccination knowledge and awareness

To assess the level of awareness, we asked respondents whether they have ever heard of a vaccine, whether they know which vaccines expectant women receive and whether they have ever heard of tetanus toxoid vaccine. Almost all respondents, 98% (102/104), were aware of vaccines Figure 1A but only 70% (73/104) knew which vaccine women receive when expectant Figure 1B. Ironically 93% (97/104) reported knowing the existence of tetanus vaccine Figure 1C.

**Figure 1.**
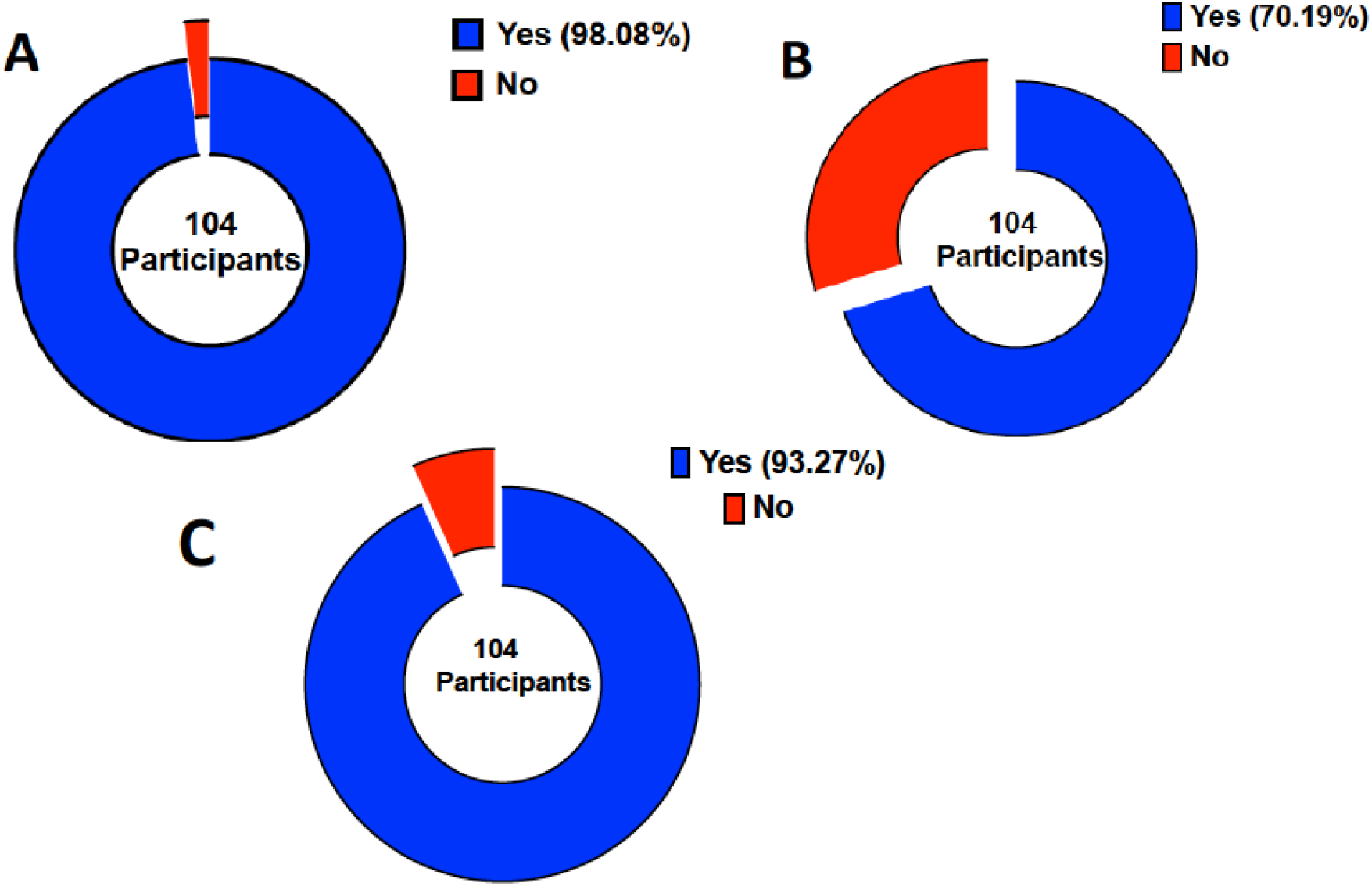
Maternal vaccine knowledge and awareness. A) Have you ever heard of vaccines? B) Do you know which vaccines women get when they are expectant? C) Have you ever heard of tetanus vaccine?

Expectant mothers reported that they are informed by the healthcare worker about the vaccine they received, although they don’t interrogate what the vaccine does. In an FGD, the following was captured:

> *“When you are told you are being given the tetanus injection, you know it protects both you and the child concerning tetanus so we don’t ask questions with regard to its purpose…We are not usually told why we are getting the injection.” Expectant mother FGD4*

Health care workers agree with the mothers, that they indeed are aware that they should receive a vaccine. Sometimes the mothers themselves can inquire why their counterparts were receiving the vaccine and not them. As per the Kenya Expended Programme for Immunization schedule mothers are supposed to get the vaccine according to their number of pregnancies. What the mothers aren’t aware of is the importance of receiving the vaccine as captured in the excerpt:

> *“I could say that majority of the mothers are aware that they are supposed to get immunization when they are expectant…As far as the knowledge of the vaccine is low in terms of knowing like what the vaccine does, why they are being immunized and what it entails.” Key informant 01*

### Reasons for not taking maternal tetanus toxoid vaccine

Out of 104 participants, 26% reported that they knew people in their community who have ever refused a vaccine when expectant Figure 2A and 8% had ever heard of negative information to vaccines in the recent past in general Figure 2B.

**Figure 2.**
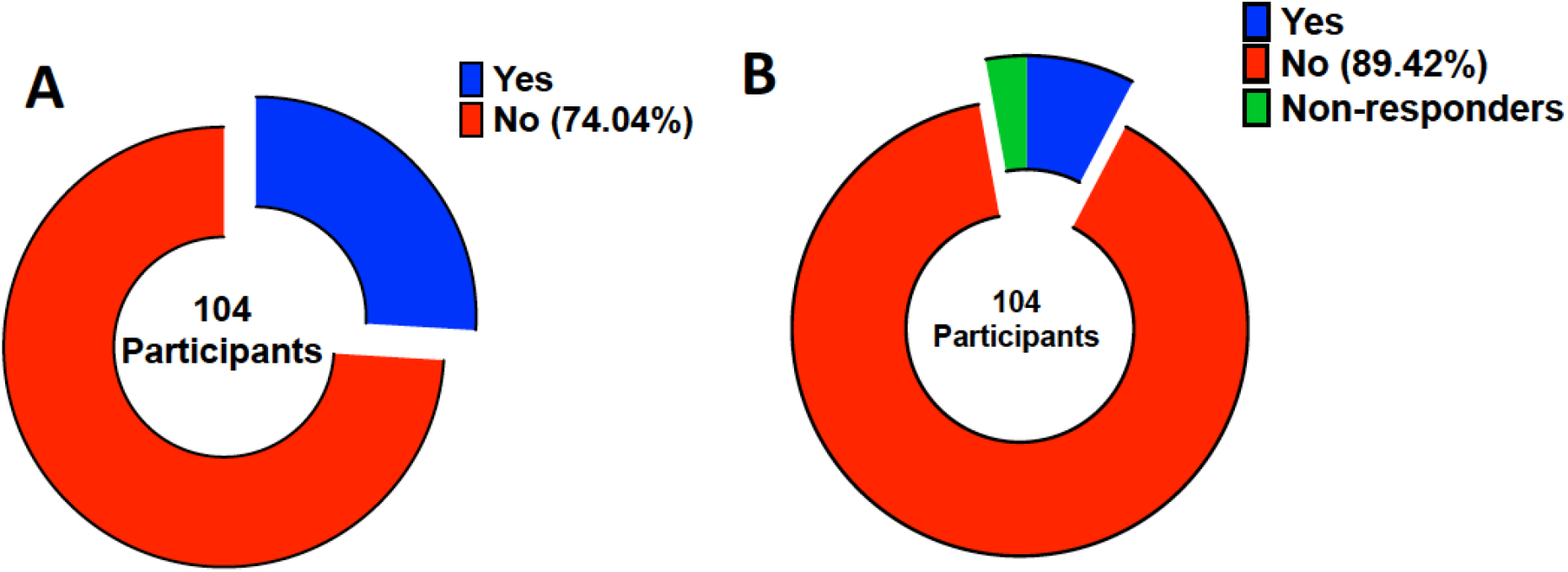
Refusal to take a vaccine and negative news about vaccines. A) Do you know people in your community who have refused or are hesitant to take a vaccine when expectant? B) Have you heard in the past negative information about vaccines?

Some of the reasons given include rumors that the tetanus vaccine is for family planning. This is captured aptly in an FGD as follows:

> *“You hear people say that they are for family planning, people say that they reduce your fertility.” Expectant Mother FGD4*

The study was also informed that others miss to take the vaccine because of home deliveries. This emerged in an FGD that:

> *“There is a neighbor who was expectant from home…For all the 9 months she has been home…She even delivered at home recently.” Expectant Mother FGD2*

Another reason was mentioned as fear of being tested for Human Immunodeficiency Virus (HIV) as evidence in the voice that:

> *“Getting tested (for HIV) is why they fear coming to clinic…Because when you come, you cannot get services until your HIV status is known.” Expectant Mother FGD1*

The respondents also adduced religious reasons. Indeed, several studies have examined whether religion influences childhood immunization uptake (Fournet et al. 2018; Costa et al. 2020) including the adolescent human papillomavirus (Best et al. 2019). Within our setting, participants reported that people from particular religious groups do not subscribe to vaccine uptake when expectant or even for their children. In particular, the study observed that:

> *“There are those who don’t come to the clinic at all and they have other reasons like religion…There is a church that doesn’t believe in hospital and its services…They say God will help…They conceive and get to delivering without any vaccinations…God keeps them safe…They exist but are not so many, others are in that sect but still come for clinic…But those who hold on to that faith, don’t come totally.” Key informant 002*

### Role of decision-makers within the household

The decision on whether a young mother will seek vaccination is partly informed by the advice she will receive from decision makers in the homestead. At least within our settings, a mother-in-law or older women have a role in influencing health care services including vaccination. As captured in the except below:

> *“There are those old women, our grandmothers who in their time did not go to the hospital or anywhere when they became expectant, even in delivery they got assistance from village Traditional Birth Attendants (TBAs) so as a daughter in-law when you marry into that home, it is not easy to convince this woman to go to hospital because they say I gave birth to your husband and I did not go to the hospital and I gave birth without any issues and right now he has grown without any problems, he is okay till now he has married you, so don’t stress yourself, just stay here I will look after you or a friend of mine will look after you.” Expectant mother FGD1*

### Trust in healthcare workers and the government

The wellcome.org/reports/wellcome-global-monitor/2018 (Wellcome 2019) the world’s largest study into how people around the world think and feel about science and major health challenges, estimates that globally 73% of people highly trust health care workers for their sources of information. We asked participants to rank from a list of healthcare workers, family or other relative, radio/TV and friends which is their most trusted source of information about vaccines for expectant mothers. Majority of the participants ranked healthcare workers as their most trusted source of information, Figure 3.

**Figure 3.**
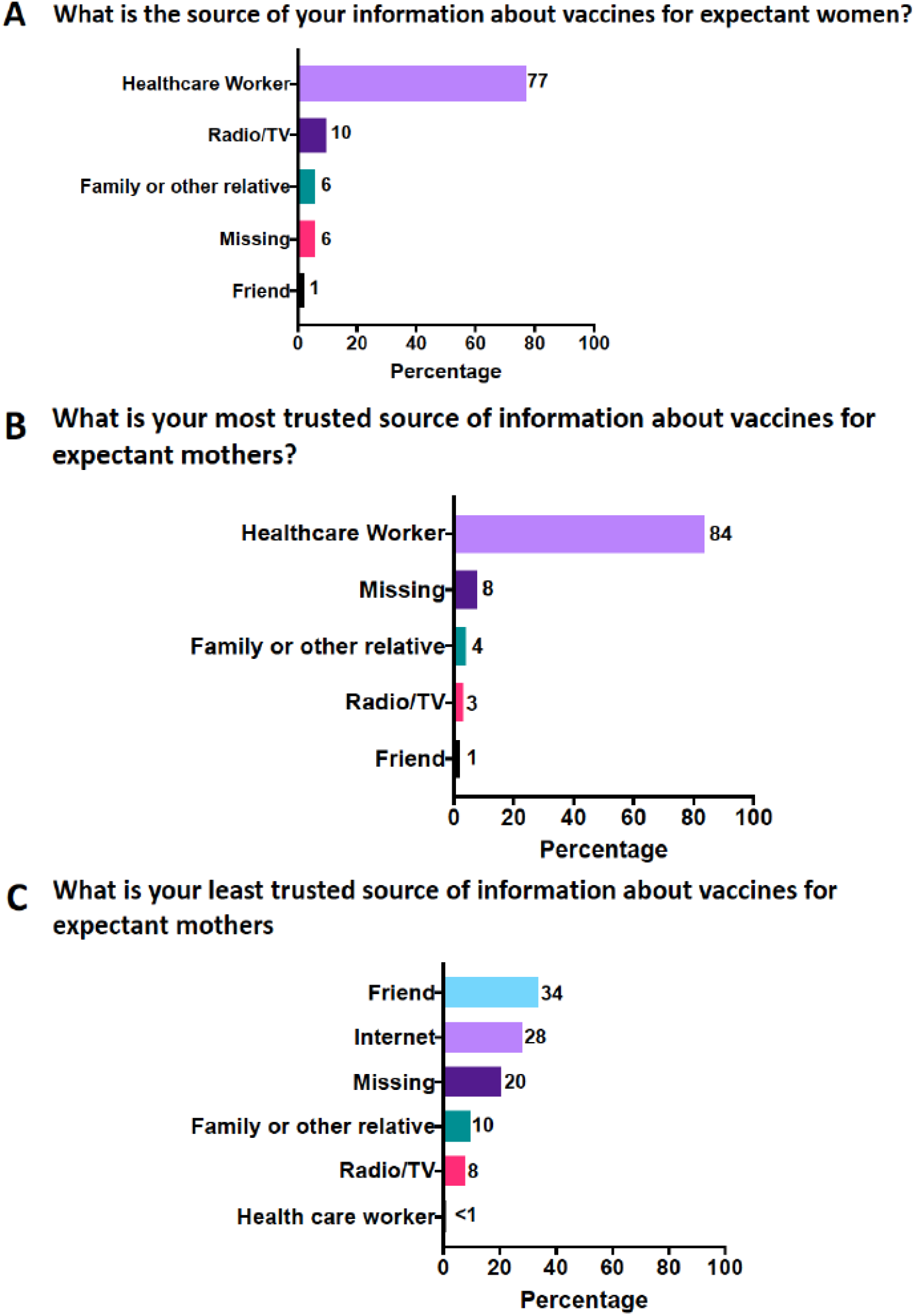
Participants (n=104) response on which is their source, most trusted and least trusted source for information about vaccines for expectant mothers

As the healthcare workers are the frontline government service providers, we tested the level of trust with the government and whether participants thought vaccines should be compulsory. Approximately 77% and 20% of the participants strongly agree or agree respectively that they trust that the government is making a decision in their best interest with regards to vaccines for expectant mothers. In contrast 53% and 23% of the participants strongly agree or agree that vaccines for expectant mothers should be made compulsory respectively.

### Health systems factors

Among the key informants, one of the reasons maternal vaccine uptake is low is vaccine stockout, which affects vaccine delivery. One key informant told the study that:

> *“At times we have get stock outs … yes as much as they (expectant mothers) would wish to get the vaccine by the time they come the vaccine is not there. Key informant 4*

The attitude of the healthcare worker also plays a role on whether a mother will come back to take the vaccine or even visit the antenatal clinic as revealed by a key informant that:

> *“Another thing which will make these mothers not to come is the attitude of the health provider at the facility if a mother comes every day and am like you are shouting at me you are not giving me attention, it is like I tell you are not listening to me you are so arrogant I will not come back I would rather stay at home”. Key informant 6*

Healthcare workload was noted as affecting vaccine delivery, especially in the lower-level health facilities whose main responsibility is to administer vaccines and offer other ANC services. A respondent had this to say:

> *“Majority of our dispensaries are run by only one nurse and only one staff…running a facility you are the one giving immunizations, you are the one taking care of the pregnancy, you are the one taking care of the sick ones who are coming in, so it was actually hectic and by the end of the day they also need to rest … come at night, they cannot work day and night, at night they are home … and over the weekends again they are taking their offs.” Key informant 5*

Since 2013, the Kenyan government introduced two policies to reduce maternal and neonatal deaths: abolition of client user fees and provision of free maternity services in public health facilities (Pyone et al. 2017). We, therefore, tested to what extent other factors such as cost in terms of fare to the clinic, waiting time at the clinic, time needed to get to the clinic, timing of the clinic and distance to the clinic prevented expectant mothers from going to get the vaccine. In all instances, these factors affected vaccine uptake only to some extent Figure 4.

**Figure 4.**
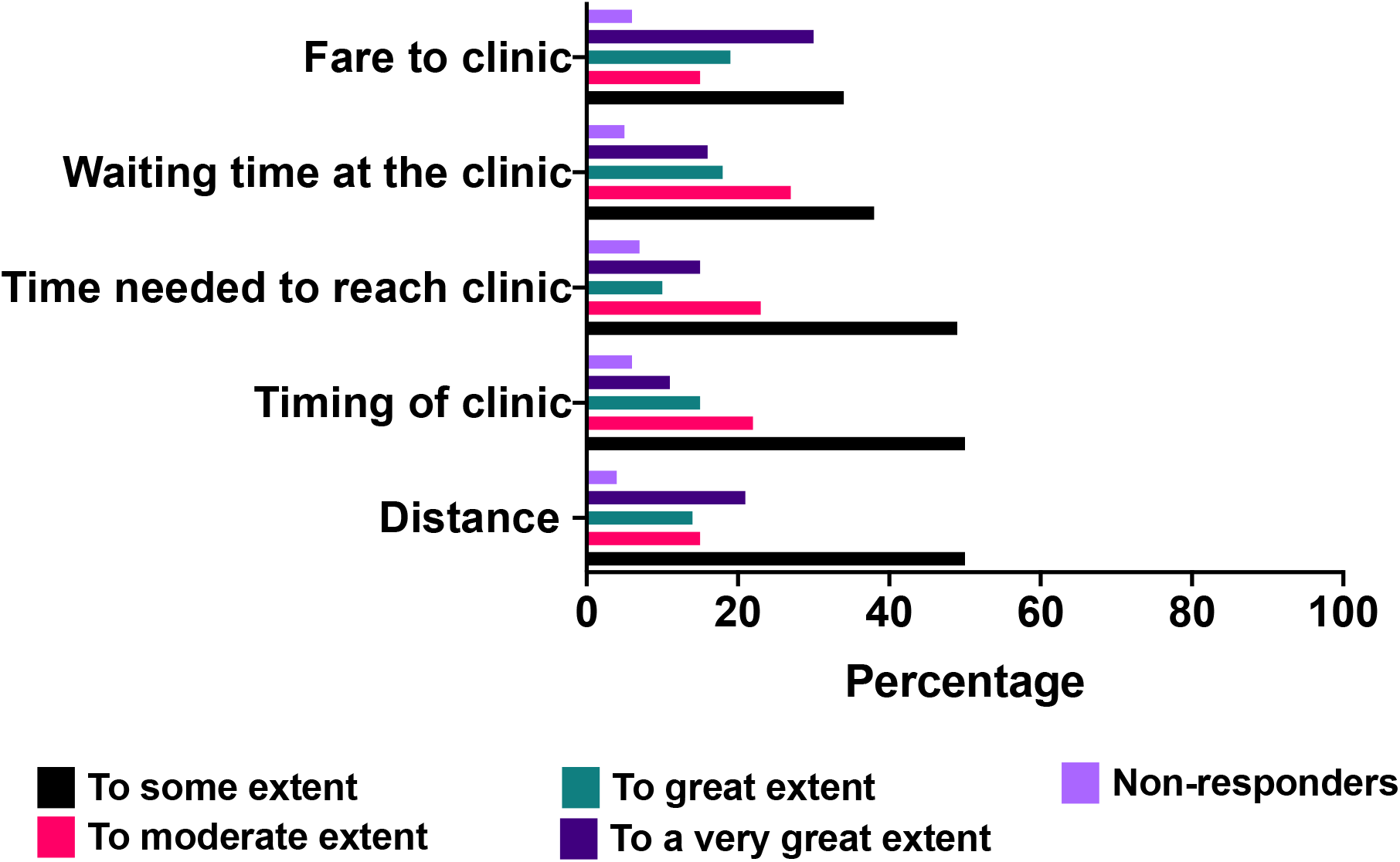
Barriers to maternal tetanus toxoid vaccine uptake

## Discussion

It is well accepted that maternal vaccination significantly reduces neonatal morbidity and mortality (Blencowe et al. 2010). However, a growing number of people who perceive vaccines as unsafe and unnecessary (Omer et al. 2009). In this study we sought to understand factors that influence maternal vaccine uptake. We report results from the contextual influences and individual/social group influences.

Almost everyone reported general knowledge of the existence of vaccines. However, few knew how the tetanus toxoid maternal vaccine works and why it is administered to mothers. The lack of knowledge can be partly explained by the low education levels of our study participants; most of them had primary level education. It is important to note that participants generally agreed that the tetanus vaccine was good for the mothers. Several factors limited maternal vaccine uptake. Rumors and misinformation around the tetanus vaccine within the community caused fears among the expectant mothers. It was perceived that the tetanus vaccine was a family planning method, a message which was reinformed when the Kenyan government rolled out tetanus vaccination campaign among school going girls. The fact that boys were excluded in the campaign made the community suspicious that the vaccine was to control fertility among young women. The majority of the Kenyans identify themselves with a particular religion. Indeed, in this study, 98% of the participants were Christians. We found that particular religious groups or sects were against modern medicine and particularly advised their congregants against using family planning methods. Additionally, they made those who heard the rumors about tetanus vaccine as a form of birth control to avoid going for the vaccine. It is worthwhile to note that the government through the public health department has put in place mechanisms to trace those missing out on vaccination and sometimes using the law to encourage vaccine uptake.

Although there has been concerted efforts by the Kenyan government to increase hospital access by rural mothers, a study conducted by Moindi et al., 2015 exploring risk perceptions associated with home deliveries estimated that approximately 26% of mothers in Kilifi County delivered at home (Moindi et al. 2016). We found that Traditional Birth Attendants (TBAs) remained important because mothers could access them easily and faster, especially if the dispensaries were far. Some mothers only sought the services of these TBAs throughout their pregnancy and as a result miss out on vaccinations. Within our setting, health care seeking behaviors are partly influenced by older women within the homestead. We found that if these older women did not go for ANC clinics, they tended to discourage young mothers married to their sons from going to the clinic for vaccines when expectant.

Global trends indicate that healthcare workers remain the most trusted sources of information on health matters (Wellcome 2019). In this study, almost 80% of the participants said they could consult healthcare worker on information related to vaccines. Very few people searched the internet if they had issues with vaccine. Considering this is a predominantly rural population, the cost of internet and smartphones could be a limiting factor.

Health systems factors such as vaccine stockout was also identified by the key informants as a challenge to vaccine delivery. As much as the healthcare workers would like to maintain a high vaccine coverage, issues such as delays in vaccine delivery to the health facilities meant that even when mothers showed up and demanded to be vaccinated the vaccines were out of stock, causing a missed opportunity. Similar to what has been found in Ethiopia (Gebremichael et al. 2018), Tanzania (Maluka et al. 2020) and Uganda (Kajungu et al. 2020), we observed that if a health care worker was harsh, abusive or rude, it could discourage women from attending ANC clinics, which has a ripple effect on uptake of vaccines by the mothers.

### Study limitations

Majority of our study populations were drawn from a rural area. It has been shown that rural areas tend to have lower education and health literacy (Das et al., 2017), factors that are important for vaccine uptake (Lorini et al., 2018). Therefore, our findings need to be interpreted within the context in which the study was done. Future studies are needed to explore in-depth maternal vaccine hesitancy issues in urban areas where there is varying education levels and social economic status.

## Conclusion

Our study highlights factors that contribute to low maternal vaccine uptake. Continuous community engagement is required to address the fears, misconceptions and rumors around tetanus vaccine uptake, important for laying grounds for future vaccine rollout for expectant mothers.

## Data Availability

We collected baseline characteristics from the study participants which is presented in Table 1 as count and percentage. Qualitative data are presented as quotes in the main text.

## Acknowledgements

This research would not have been possible without the immense support of Kilifi County Department of Health, Community Health Volunteers, local chiefs and community members who participated in the study. We thank Pwani University for the scientific and ethical review. This manuscript was submitted for publication with the permission of the Director KEMRI

